# Association of Childhood Exposure to School Racial Segregation with Late-Life Cognitive Outcomes among Older Americans

**DOI:** 10.1101/2024.06.21.24309186

**Authors:** Zhuoer Lin, Yi Wang, Thomas M. Gill, Xi Chen

## Abstract

**IMPORTANCE:** Disparities in cognition, including dementia occurrence, persist between White and Black older adults, and are possibly influenced by early educational differences stemming from structural racism. However, the relationship between school racial segregation and later-life cognition remains underexplored.

**OBJECTIVE:** To investigate the association between childhood contextual exposure to school racial segregation and cognitive outcomes in later life.

**DESIGN, SETTING, AND PARTICIPANTS:** Data from 16,625 non-Hispanic White (hereafter, White) and 3,335 non-Hispanic Black (hereafter, Black) Americans aged 65 or older were analyzed from the Health and Retirement Study.

**EXPOSURES:** State-level White-Black dissimilarity index for public elementary schools in the late 1960s (range: 0-100) was used to measure school segregation. States were categorized into high segregation (≥83.6) and low segregation (<83.6) based on the top quintile.

**MAIN OUTCOMES AND MEASURES:** Cognitive scores, cognitive impairment (with or without dementia), and dementia were assessed using the Telephone Interview for Cognitive Status (TICS) and proxy assessment. Multilevel regression analyses were conducted, adjusting for demographic covariates, socioeconomic status, and health factors. Stratified analyses by race were performed.

**RESULTS:** The mean (SD) age of participants was 78.5 (5.7) years, and 11,208 (56.2%) were female. Participants exposed to high segregation exhibited lower cognitive scores (12.6 vs. 13.6; *P*<0.001) and higher prevalence of cognitive impairment (50.8% vs 41.4%; *P*<0.001) and dementia (26.0% vs. 19.5%; *P*<0.001), compared to those with low segregation exposure. Multilevel analyses revealed a significant negative association between school segregation and later-life cognitive even after adjusting sequentially for potential confounders, and these associations were stronger among Black than White participants. Notably, in the fully adjusted model, Black participants exposed to high segregation displayed significantly lower cognitive scores (−0.51; 95% CI: −0.94, −0.09) and higher likelihood of cognitive impairment (adjusted Odds Ratio [aOR]: 1.45, 95% CI: 1.22, 1.72) and dementia (aOR: 1.31, 95% CI: 1.06, 1.63).

**CONCLUSIONS AND RELEVANCE:** Our study underscores that childhood exposure to state-level school segregation is associated with late-life cognition, especially for Black Americans. Given the rising trend of school segregation in the US, educational policies aimed at reducing segregation are crucial to address health inequities. Clinicians can leverage patients’ early-life educational circumstances to promote screening, prevention, and management of cognitive disorders.

**Key Points:** *Questions:* Is state-level school racial segregation during childhood associated with cognitive outcomes in later life among non-Hispanic Black (hereafter, Black) and non-Hispanic White (hereafter, White) Americans?

*Findings:* In this nationally representative sample, older adults exposed to high levels of school segregation had lower cognitive scores and an increased likelihood of cognitive impairment and dementia compared to those with low levels of exposure. These associations remained significant after adjusting for a comprehensive array of factors over the life course, and were more pronounced for Black than White participants.

*Meaning:* These findings suggest that investments to reduce school racial segregation could have lasting benefits for cognition and racial equity, spotlighting school racial segregation as an important form of structural racism in the US. Ascertainment of school racial segregation during childhood could help to promote more efficient and equitable screening, prevention, and management of cognitive impairment in clinical settings.

## Introduction

Cognitive impairment poses considerable challenges for older persons,^1^ with Alzheimer’s disease and related dementias affecting millions of Americans and the burden escalating as the population ages. Notably, marked racial and ethnic disparities persist.^2^ Cognitive disorders disproportionally impact disadvantaged populations, diminishing individual well-being and imposing substantial burdens on caregivers and families, thereby exacerbating societal racial and ethnic disparities.^3^

Emerging evidence underscores the profound influence of adverse early-life circumstances on brain development, which can lead to declines in cognitive function over the lifespan.^4,5,6^ Racial differences in early educational environments, particularly those rooted in structural racism, appear pivotal in shaping cognition in later life.^7–9^

School racial segregation (school segregation hereafter), a significant aspect of United States (US) education systems, may exert a particularly profound impact on cognition.^10^ This practice physically segregates students in educational institutions based on racial backgrounds, resulting in vastly unequal educational experiences, qualities, and opportunities between White and minoritized populations. Despite the historic *Brown v. Board* ruling, US schools continue to struggle with heightened levels of segregation,^11,12^ with more than half of students attending schools in districts that are predominantly white or nonwhite, and approximately 40% of black students attending schools that are 90% to 100% nonwhite.^13,14^

Understanding the long-term relationship between school segregation and later-life cognition is imperative, as the school environment not only shapes educational outcomes but also influences the overall quality of the educational experience.^15^ Disadvantaged educational experiences and outcomes may subsequently shape cognitive development and exert enduring influences on cognition later in life. Moreover, it is vital to differentiate the impacts of school segregation on cognitive impairment measures that vary in severity. Effects on non-Hispanic Black (hereafter, Black) and non-Hispanic White (hereafter, White) Americans may also fundamentally differ, as attending segregated schools can predominantly expose Black children to discrimination, racism, diminished school resources, and childhood adversities.^16,17^

Previous studies evaluating the association between US school segregation and health outcomes in later life have been limited by a singular focus on indirect measures of school segregation,^18^ reliance solely on self-reported data,^19–21^ lack of nationally representative samples,^19,22,23^ and inattention to later-life cognitive outcomes.^24–26^ To bridge these research gaps, our study assesses whether and how childhood contextual exposure to school segregation is associated with cognitive outcomes in later life. We linked historical data on White-Black school segregation in public elementary schools from the late 1960s to a nationally representative sample of older adults in the US, utilizing the Health and Retirement Study (HRS), which includes rigorous cognitive assessments and a comprehensive array of sociodemographic and health factors over the life course. We hypothesized that childhood exposure to high levels of school segregation is associated with poorer later-life cognitive outcomes, especially among Black older adults.

## Methods

This study was exempt from full review by the Yale University ethics review committee, with informed consent waived due to the use of deidentified and publicly available data (IRB Protocol ID: 2000031713). The study adhered to the Strengthening the Reporting of Observational Studies in Epidemiology (STROBE) reporting guideline.

### Data and Study Participants

Data for this study were derived from two primary sources: 1) school segregation data using public elementary school enrollment and segregation information from the Office of Civil Rights (OCR) of the US Department of Health and Human Services; and 2) population-based survey data from the HRS in the US.^27,28^

The school segregation data were obtained from OCR files that included school enrollment statistics and segregation index for school districts in the US, stratified by non-Hispanic Black (hereafter, Black), non-Hispanic White (hereafter, White), Hispanic and Asian populations. Previous studies have thoroughly cleaned and validated the OCR data, constructing segregation index measures at the metropolitan level from the late 1960s.^27,28^ These metropolitan-level public elementary school enrollment and segregation index data, spanning 328 US metropolitan areas, were used in this study to construct within-metropolitan school segregation index for each state.^28^

The HRS is a nationally representative longitudinal survey of Americans aged 50 years and older, conducted biannually from since 1992. Each wave of the survey included approximately 19,000 participants, with consistent collection of data on cognition and individual-level sociodemographic and health characteristics since 1995. For our study, data were drawn from HRS participants surveyed between 1995 and 2018 (i.e., the most recent wave pre-COVID^29^). Given our focus on Black-White school segregation, our analysis centered on participants self-identified as non-Hispanic Black (hereafter, Black), and non-Hispanic White (hereafter, White) older adults. We analyzed their most recent cognitive assessments, specifically those closest to 2018 (pre-COVID), along with corresponding sociodemographic and health factors obtained from the same HRS survey.

The sample selection process is shown in Figure 1. Over the study period (1995-2018), 39,958 HRS participants underwent cognitive assessments. Among these, 22,798 participants were aged 65 or older, and have a valid U.S. childhood state of residence with linked data on school segregation measures. After excluding 1,441 participants without complete data on demographics, socioeconomic status (SES), or later-life health factors, as well as 1,397 participants who self-identified as Hispanic or another racial/ethnic group other than Black or White, the final sample comprised 19,960 participants aged 65 or older with complete data and measurements. This sample included 16,625 White and 3,335 Black participants.

**Figure 1.**
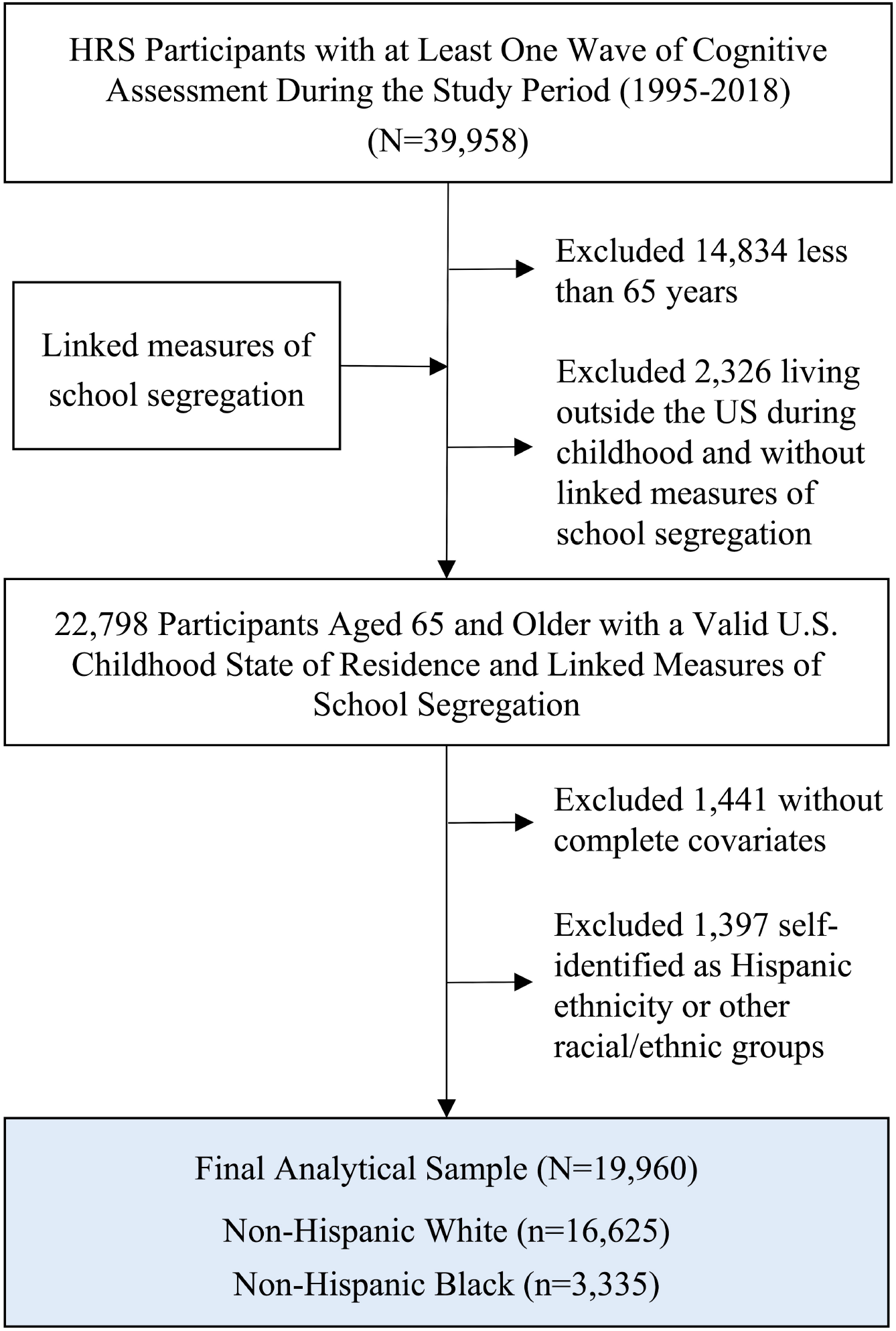
Flow chart of sample selection process *Notes*: HRS=Health and Retirement Study.

### Cognitive Outcomes

Cognition was assessed using the Telephone Interview for Cognitive Status, a 27-point cognitive scale encompassing a series of cognitive questions evaluating memory (immediate and delayed word recall, scored 0-20 points), working memory (serial sevens substation tests, scored 0-5 points), and speed of mental processing (counting backwards test, scored 0-2 points). The scale reflects global cognitive function, with higher score indicating better cognitive performance. Cognitive impairment and dementia status were determined based on established criteria: a score below 12 indicated cognitive impairment (with or without dementia), and a score below 7 indicated dementia.^30,31^

For participants unable to complete the cognitive assessment by themselves, a 11-point proxy cognitive scale was constructed to determine cognitive status. This scale incorporated proxy assessment of memory, limitations in instrumental activities of daily living (IADL), and interviewer assessments of cognitive impairment, with a higher score indicating poorer cognitive function. Cognitive impairment (with or without dementia) was determined if the proxy score was above 2, and dementia status was identified if the proxy score was above 5.^30,31^

All study participants (16,231 White and 3,235 Black) had valid assessments of cognitive impairment (dichotomous: 0/1) and dementia (dichotomous: 0/1) in their latest wave. Among them, 13,394 White and 2,596 Black participants had complete data on the 27-point cognitive scale (continuous: 0-27).

### School Segregation

School segregation was assessed using the White-Black dissimilarity index (hereafter referred to as the dissimilarity index), which measures the extent of segregation between Black and White students in public elementary schools within metropolitan areas. Scores on this index, ranging from 0 to 100, indicate the percentage of Black children who would need to move to a different school to achieve an equal distribution of Black and White students across schools in a metropolitan area. Higher scores indicate greater levels of segregation. The dissimilarity index primarily relied on enrollment data from public elementary schools reported by the OCR in 1968, supplemented by data from 1969 to 1971.^27,28^

Because all study participants attended elementary schools prior to 1970 (i.e. before significant desegregation efforts were implemented from the 1970s to the 1990s), using late 1960s data for calculating the dissimilarity index is highly relevant.^28,32^ Participants were asked to report the state where they lived when they were 10 years old. Subsequently, an average dissimilarity index score was calculated for each corresponding state during the late 1960s, weighted by the enrollment numbers of Black and White students in public elementary schools within the respective metropolitan areas.

Due to the skewed distribution of index scores (Supplementary eFigure 1), states were categorized based on quintile of scores, a commonly used cut-off in prior contextual-level research in older Americans.^33^ States in the highest quintile (dissimilarity index ≥ 83.6) were classified as “high segregation”, while the others were classified as “low segregation”.

### Covariates

Age, sex, and race were included as the demographic covariates. Additionally, we incorporated a comprehensive array of socioeconomic status (SES) and health factors, recognized as the key risk factors for cognitive impairment and dementia over the life course.^6,34^ Given merging evidence suggesting associations between school segregation and SES and health, it is important to account for these factors as they may serve as pivotal links between school segregation and cognition.^5,22,35,36^

### Statistical Analyses

Descriptive statistics were estimated for the entire sample, as well as for subgroups with high versus low levels of segregation. Differences in characteristics were assessed using appropriate statistical tests: Chi-square tests for categorical variables, Welch t-tests for continuous variables, and Mann-Whitney-Wilcoxon tests for ordinal variables.

To evaluate the association between childhood school segregation exposure and cognitive outcomes in later life, multilevel models were employed with participants as level 1 and childhood states of residence as level 2. Random intercepts for each childhood states of residence were included in the model to address unobserved heterogeneity and differences between states. Robust standard errors, clustered at the state level, were estimated to account for within-state correlation.^37,38^

In a set of hierarchical models, we sequentially adjusted for factors over the life course that could be associated with school segregation and subsequently affect cognitive outcomes. In Model A, we adjusted solely for demographic factors; in Model B, mid-to-late life SES factors (e.g., years of education, wealth level, insurance coverage) were added; and in Model C, late-life health factors (e.g., hypertension, diabetes, smoking) were added. Linear models were used for the continuous outcome (i.e., cognitive score) and logit models were used for the dichotomous outcomes (i.e., cognitive impairment and dementia). Given the inherent heterogeneity in the association of school segregation by race, models were estimated separately for Black and White participants.

All analyses were performed using Stata 17.0 (StataCorp LLC), and all tests were two-sided with statistical significance set at *P*<0.05.

## Results

### Sample Characteristics

The sample characteristics are presented in Table 1. Among the 19,960 participants, 5,066 resided in states characterized by high segregation, while 14,894 lived in states with low segregation. The participants’ mean (SD) level of school segregation, as indicated by the dissimilarity index, was 80.0 (5.7); and the level was significantly higher for those who lived in high segregation states (mean: 86.9) than low segregation states (mean: 77.6) (*P*<0.001). Participants exposed to high segregation during childhood had significantly lower cognitive scores (12.6 vs 13.6; *P*<0.001), along with a higher likelihood of cognitive impairment (50.8% vs. 41.4%; *P*<0.001) and dementia (26.0% vs 19.5%; *P*<0.001) compared to their counterparts with low segregation exposure. As shown in Supplementary eFigure 2, participants with high segregation demonstrated consistently poorer cognitive outcomes across various ages when compared to those with low segregation exposure (Supplementary eFigure 2).

**Figure 2.**
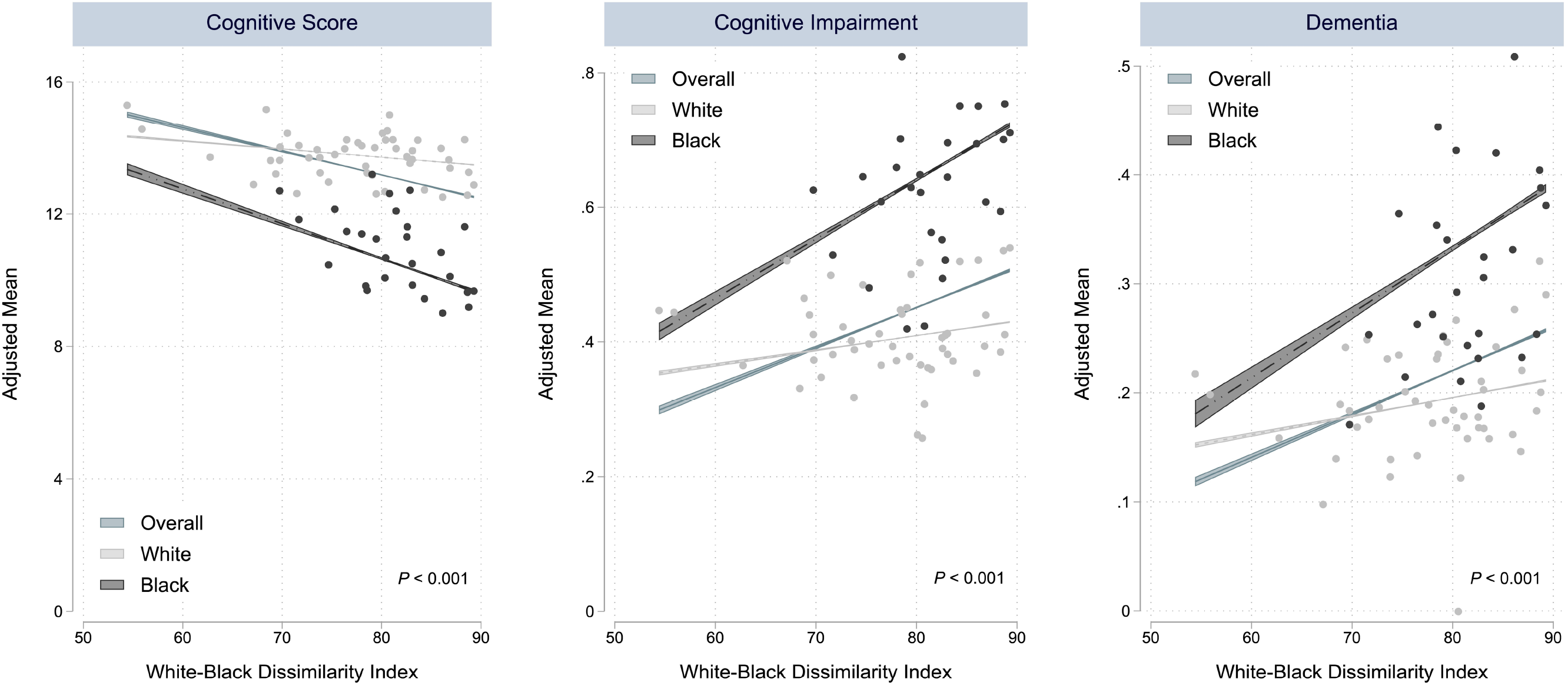
Relationship between school segregation and cognitive outcomes by race *Notes*: The figure presents scatterplots of US states demonstrating the inverse relationships between school segregation (measured by White-Black dissimilarity index) and cognitive function, with dissimilarity index on the x-axis and adjusted cognitive outcomes on the y-axis. The scatterplots are stratified by White (in gray color) and Black (in black color) participants. White refers to non-Hispanic White, and Black refers to non-Hispanic Black. The average cognitive outcomes were estimated respectively for White and Black participants in each state after adjusting for age and sex; and only states with more than 10 observations are plotted. The fitted lines (with 95% CI) denote the linear relationship between school segregation and adjusted average cognitive outcomes for all participants (in blue color), White participants (in gray color), and Black participants (in black color). Chow cross-equation tests were performed to denote if there were statistically significant differences in fitted slopes between White and Black participants for each cognitive outcome, and *P*-values were displayed at the bottom of the corresponding panel.

**Table 1.** Characteristics of study participants by school segregation.

|  | Overall<br>(N=19,960) | Low<br>Segregation<br>(N=14,894) | High<br>Segregation<br>(N=5,066) | *P*-values <sup>a</sup> |
| --- | --- | --- | --- | --- |
| **School Segregation** |  |  |  |  |
| Dissimilarity Index (0-100), mean (SD) | 80.0 (5.7) | 77.6 (4.5) | 86.9 (1.9) | <0.001 |
| **Cognitive Outcomes** |  |  |  |  |
| Cognitive Score (0-27), mean (SD) | 13.3 (5.0) | 13.6 (4.9) | 12.6 (5.2) | <0.001 |
| Cognitive Impairment, No. (%) | 8735 (43.8) | 6163 (41.4) | 2572 (50.8) | <0.001 |
| Dementia, No. (%) | 4216 (21.1) | 2899 (19.5) | 1317 (26.0) | <0.001 |
| **Demographic Covariates** |  |  |  |  |
| Age, mean (SD) | 78.5 (8.4) | 78.5 (8.4) | 78.5 (8.5) | 0.887 |
| Female, No. (%) | 11208 (56.2) | 8321 (55.9) | 2887 (57.0) | 0.165 |
| Race |  |  |  | <0.001 |
| Non-Hispanic White, No. (%) | 16625 (83.3) | 13000 (87.3) | 3625 (71.6) |  |
| Non-Hispanic Black, No. (%) | 3335 (16.7) | 1894 (12.7) | 1441 (28.4) |  |
| **Socioeconomic Status (SES)** |  |  |  |  |
| Years of Education, mean (SD) | 12.3 (3.0) | 12.6 (2.8) | 11.7 (3.5) | <0.001 |
| Wealth Level |  |  |  | <0.001 |
| Lowest, No. (%) | 4934 (24.7) | 3466 (23.3) | 1468 (29.0) |  |
| Lower-middle, No. (%) | 4707 (23.6) | 3390 (22.8) | 1317 (26.0) |  |
| Upper-middle, No. (%) | 5100 (25.6) | 3910 (26.3) | 1190 (23.5) |  |
| Highest, No. (%) | 5219 (26.1) | 4128 (27.7) | 1091 (21.5) |  |
| Medicare Enrollment, No. (%) | 19309 (96.7) | 14397 (96.7) | 4912 (97.0) | 0.304 |
| Medicaid Enrollment, No. (%) | 2489 (12.5) | 1654 (11.1) | 835 (16.5) | <0.001 |
| Military Health Care Plan Enrollment, No. (%) | 1262 (6.3) | 946 (6.4) | 316 (6.2) | 0.774 |
| Private Health Insurance Coverage, No. (%) | 9513 (47.7) | 7211 (48.4) | 2302 (45.4) | <0.001 |
| Employer-based Health Insurance Coverage, No. (%) | 5047 (25.3) | 3959 (26.6) | 1088 (21.5) | <0.001 |
| **Health Factors** |  |  |  |  |
| Hypertension, No. (%) | 13807 (69.2) | 10168 (68.3) | 3639 (71.8) | <0.001 |
| Diabetes, No. (%) | 5402 (27.1) | 3964 (26.6) | 1438 (28.4) | 0.014 |
| Heart Disease, No. (%) | 8562 (42.9) | 6377 (42.8) | 2185 (43.1) | 0.696 |
| Stroke, No. (%) | 3808 (19.1) | 2811 (18.9) | 997 (19.7) | 0.207 |
| Obesity, No. (%) | 12312 (61.7) | 9145 (61.4) | 3167 (62.5) | 0.159 |
| ADL Limitations, No. (%) | 7528 (37.7) | 5406 (36.3) | 2122 (41.9) | <0.001 |
| IADL Limitations, No. (%) | 7231 (36.2) | 5211 (35.0) | 2020 (39.9) | <0.001 |
| Currently Smoking, No. (%) | 1894 (9.5) | 1380 (9.3) | 514 (10.1) | 0.065 |
| Ever Smoking, No. (%) | 11775 (59.0) | 8926 (59.9) | 2849 (56.2) | <0.001 |

Abbreviations: SD=standard deviation, IQR=interquartile range, ADL=activities of daily living, IADL=instrumental activities of daily living.
<sup>a</sup> Differences in characteristics were assessed using appropriate statistical tests: Chi-square tests for categorical variables, Welch t-tests for continuous variables, and Mann-Whitney-Wilcoxon tests for ordinal variables.

The two segregation groups were comparable in terms of age and sex, but differed significantly according to several of the other characteristics (Table 1). Relative to low segregation states, high segregation states included a higher proportion of Black participants (28.4% vs 12.7%, *P*<0.001), a higher proportion of participants with disadvantaged SES, including lower educational attainment, decreased wealth level, higher Medicaid enrollment, and a higher proportion of participants with several health conditions, including hypertension, diabetes, ADL and IADL limitations, and a history of (ever) smoking.

### Association between School Segregation and Cognitive Outcomes

Figure 2 shows the inverse relationships between state-level dissimilarity index scores and cognition, adjusted for age and sex, across the entire sample and separately for Black and White participants. States characterized by high levels of school segregation demonstrated lower average cognitive scores and increased proportions of cognitive impairment and dementia. Notably, the slopes of the fitted lines were steeper for Black participants compared with White participants across all cognitive outcomes (*P*<0.001), indicating a stronger association of school segregation with cognitive outcomes among Black older adults.

Multilevel regression analyses, depicted in Figures 3 and 4 (with numerical estimates provided in Supplementary eTables 1-2), confirmed these trends. Overall, participants who experienced high levels of segregation during childhood exhibited notably lower cognitive scores (Figure 3) and higher odds of cognitive impairment and dementia (Figure 4) compared with those in low segregation states, after adjusting for demographic covariates (Model A). Moreover, the magnitude of coefficients was larger for Black participants than for White participants across all cognitive outcomes. Moreover, the association between school segregation and dementia for White participants was not statistically significant.

**Figure 3.**
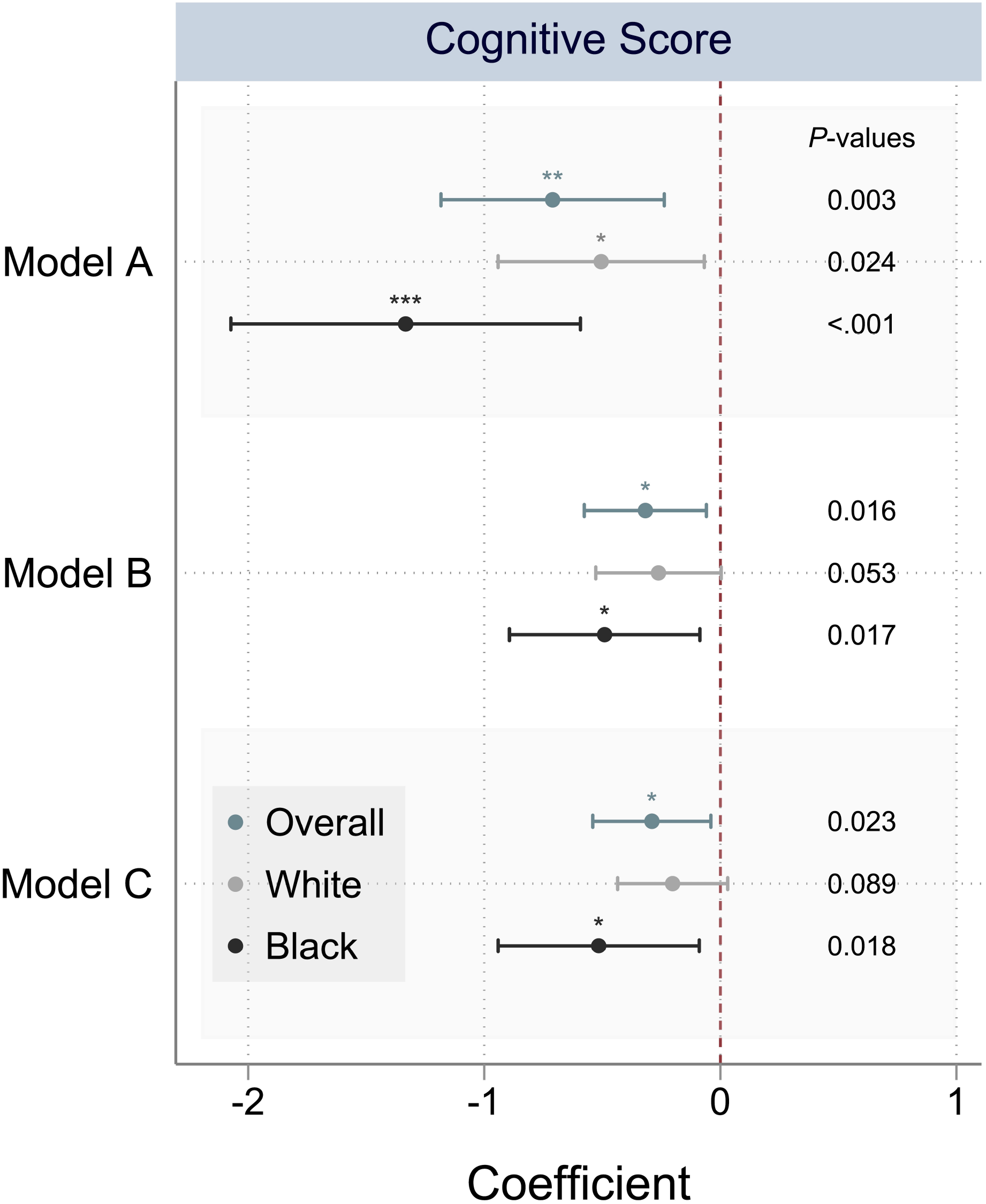
Association between school segregation and cognitive score by race *Notes*: Multilevel regression models were used to estimate the association between school segregation and cognitive score for the overall sample (in blue color) and for White (in gray color) and Black participants (in black color), respectively. White refers to non-Hispanic White, and Black refers to non-Hispanic Black. Horizontal lines represent the 95% confidence interval, and *P*-values are displayed alongside each lines to denote statistical significance. Covariates were added sequentially into the regression model. Model A adjusted for demographic covariates, including age, sex, and race. Model B additionally adjusted for life course socioeconomic status (SES), including educational attainment, wealth level, and Medicare enrollment, Medicaid enrollment, VA enrollment, private health insurance coverage, and employer-based health insurance coverage. Model C further adjusted for health factors over the life course including hypertension, diabetes, heart diseases, stroke, obesity, ADL and IADL functional limitations, and smoking behaviors. Random intercepts for each childhood states of residence were included in the models to address unobserved heterogeneity and differences between states. The numerical estimates are presented in Supplementary eTables 1-2. Asterisks denote the statistical significance of the association: *** *P* < 0.001, ** *P* < 0.01, * *P* < 0.05.

**Figure 4.**
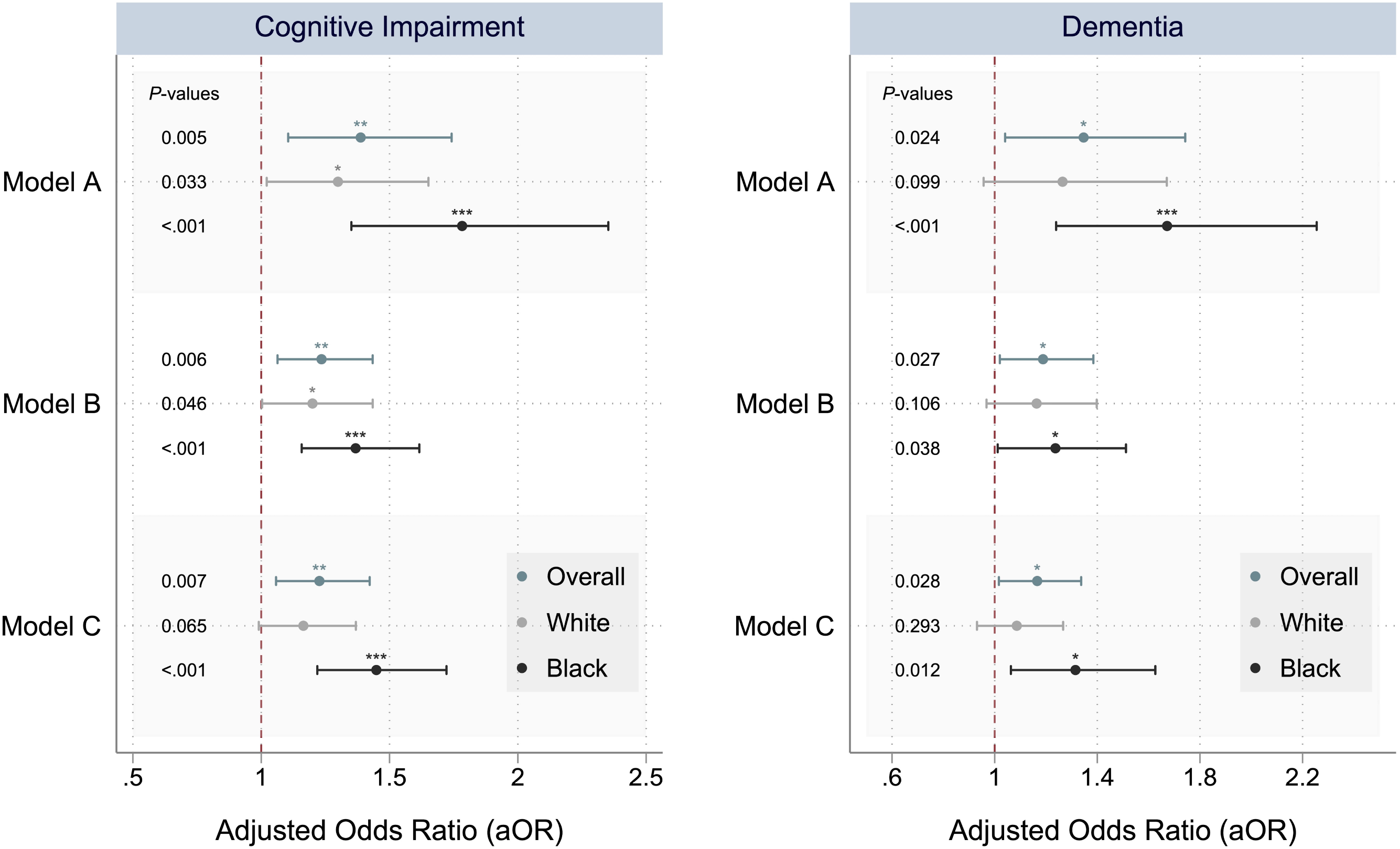
Association between school segregation and cognitive impairment and dementia by race *Notes*: Multilevel regressions were used to estimate the association between school segregation and cognitive impairment (left panel) and dementia (right panel), for overall sample (in blue color), as well as for White (in gray color) and Black participants (in black color) respectively. White refers to non-Hispanic White, and Black refers to non-Hispanic Black. Horizontal lines represent the 95% confidence interval, and *P*-values are displayed alongside each lines to denote statistical significance. Covariates were added sequentially into the regression model. Model A adjusted for demographic covariates, including age, sex, and race. Model B additionally adjusted for life course socioeconomic status (SES), including educational attainment, wealth level, and Medicare enrollment, Medicaid enrollment, VA enrollment, private health insurance coverage, and employer-based health insurance coverage. Model C further adjusted for health factors over the life course including hypertension, diabetes, heart diseases, stroke, obesity, ADL and IADL functional limitations, and smoking behaviors. Random intercepts for each childhood states of residence were included in the models to address unobserved heterogeneity and differences between states. The numerical estimates are presented in Supplementary eTables 1-2. Asterisks denote the statistical significance of the association: *** *P* < 0.001, ** *P* < 0.01, * *P* < 0.05.

The associations observed in Model A were attenuated after further adjustment for SES (Model B), but not after additional adjustment for health factors (Model C). In the fully adjusted model (Model C), participants exposed to high levels of segregation had significantly lower cognitive scores, and higher likelihood of cognitive impairment and dementia. Stratified analyses revealed that this association was more pronounced and only statistically significant among Black participants compared to White participants (Figures 3-4 and Supplementary eTables 1-2).

## Discussion

Using data from a nationally representative sample of older adults in the US, linked to historical administrative data from the late 1960s, we offer insights into the association between early life exposure to school segregation and cognitive outcomes in later life. Our findings demonstrate that childhood exposure to high levels of school segregation was associated with lower cognitive scores and a higher likelihood of cognitive impairment and dementia, particularly among Black Americans.

School segregation has long persisted within the US education system,^39,40^ yet its long-term association with health outcomes has not been thoroughly investigated due to data constraints. Our study, which utilizes a measure of school segregation derived from historical administrative records, captures contextual exposure and helps mitigate recall bias often associated with self-reported data on segregation exposure. Leveraging the data from HRS also allowed us to link these contextual segregation exposures during childhood to various robust cognitive outcomes (i.e., cognitive score, cognitive impairment, and dementia) in later-life.

To the best of our knowledge, our study is the first to employ such linked data to evaluate the association between state-level school segregation and cognitive outcomes in later life. Our results align with existing literature highlighting the negative association between adverse early-life educational experiences and cognition in later life.^9^ Notably, an increasing number of studies have indicated that higher educational quality is linked to a reduced risk of dementia and improved cognitive outcomes in later life.^41,42^ Our analysis, drawing from comprehensive national datasets, offers broader contextual insights into the relationship between school segregation and cognition, setting it apart from studies that have relied on self-reported^19–21^ or regional-specific data,^19,22^ and/or those that have not focused specifically on cognition.^24–26^

High levels of school segregation often reflect limited and unequal educational investment. States with high levels of school segregation may allocate less funding to schools serving predominantly Black students,^43^ resulting in higher teacher turnover and fewer resources for Black children compare to their White counterparts.^44,45^ This disparity not only impacts Black students’ access to education but also their educational quality and experience, potentially influencing their cognitive development and later-life cognition. Our study underscores that the negative association between school segregation and cognitive outcomes is more pronounced for Black than White participants, consistent with previous reseearch.^9,21,25,46^

Moreover, our findings suggest that SES may explain a significant portion of the association between school segregation and cognition. We observed an approximately 50% reduction in the association among Black individuals after accounting for SES, highlighting the role of SES as potential mediators linking school segregation and cognition. Prior research has linked school segregation to lower SES, manifested in lower educational attainment and subsequent earnings.^47^ As a key social determinant, SES may directly impact individuals’ cognitive development in early life and indirectly influence cognitive performance through risk factors over the life course.^6^ Factors like reduced access to healthcare and health-promoting resources,^48^ as well as the adoption of unhealthy behaviors (e.g., smoking), might play a role.^49^ Moreover, prior studies suggest that students from lower SES backgrounds tend to be more affected by their school context (e.g., student-teacher ratio, term length) compared with those from high SES backgrounds.^50^ Our findings therefore may inform the formulation of public policy and development of interventions that prioritize Black children with low SES living in highly school-segregated states.

In addition to SES, comorbidities, functional limitations, and smoking may also influence cognition in later life.^6^ Our results indicate that the association between school segregation and cognition persist even after accounting for these factors. This could be because SES already encapsulates the associations of health conditions and lifestyle choices with cognition. Future research may benefit from incorporating more nuanced measures of health and health behavior factors over the life course to elucidate the potential mechanisms.

Despite these contributions, our study has several limitations. First, the state-level measure of school segregation could dampen the actual differences. More granular administrative data linking participants to specific schools and classrooms over time, as well as data on the degree of segregation within educational settings, could reveal even more pronounced differences in exposures to school segregation. Second, the lack of data on educational quality measures, such as teacher qualifications, experience, salaries, student-teacher ratios, and duration of schooling, poses a challenge in understanding the mechanisms underlying the link between school segregation and cognition. Addressing these gaps represents a crucial area for future research. Third, we evaluated cognition at a single time point, limiting our ability to examine cognitive trajectories over time. Future research efforts should aim to explore how school segregation may be associated with both cognitive development and cognitive decline through long-term longitudinal studies. Additionally, it is important to note that our results are not causal. To enhance understanding in the area, further studies could employ quasi-experimental designs or leverage policy interventions to strengthen causal relationships.

## Conclusions

Our study reveals that childhood exposure to higher levels of school segregation is associated with poorer cognitive outcomes in older Americans, particularly among Black individuals. Our findings underscore that investments to reduce school segregation could have lasting benefits for cognition and its racial equity. By highlighting school segregation as a form of structural racism entrenched within the US, our study contributes to the broader discussion on racial disparities in cognition and underscores the imperative for addressing systemic inequities in education.

## Data Availability

All data produced in the present study are available upon reasonable request to the authors.

https://hrs.isr.umich.edu/

## Acknowledgements

This study was funded by research grant R01AG077529 from the U.S. National Institute on Aging; and grant P30AG021342 from the U.S. National Institute on Aging to the Yale Claude D. Pepper Older Americans Independence Center. The funders had no role in the study design; data collection, analysis, or interpretation; in the writing of the report; or in the decision to submit the article for publication. The authors declare no conflict of interest.

## Supplementary Online Content

**Supplementary eFigure 1.**
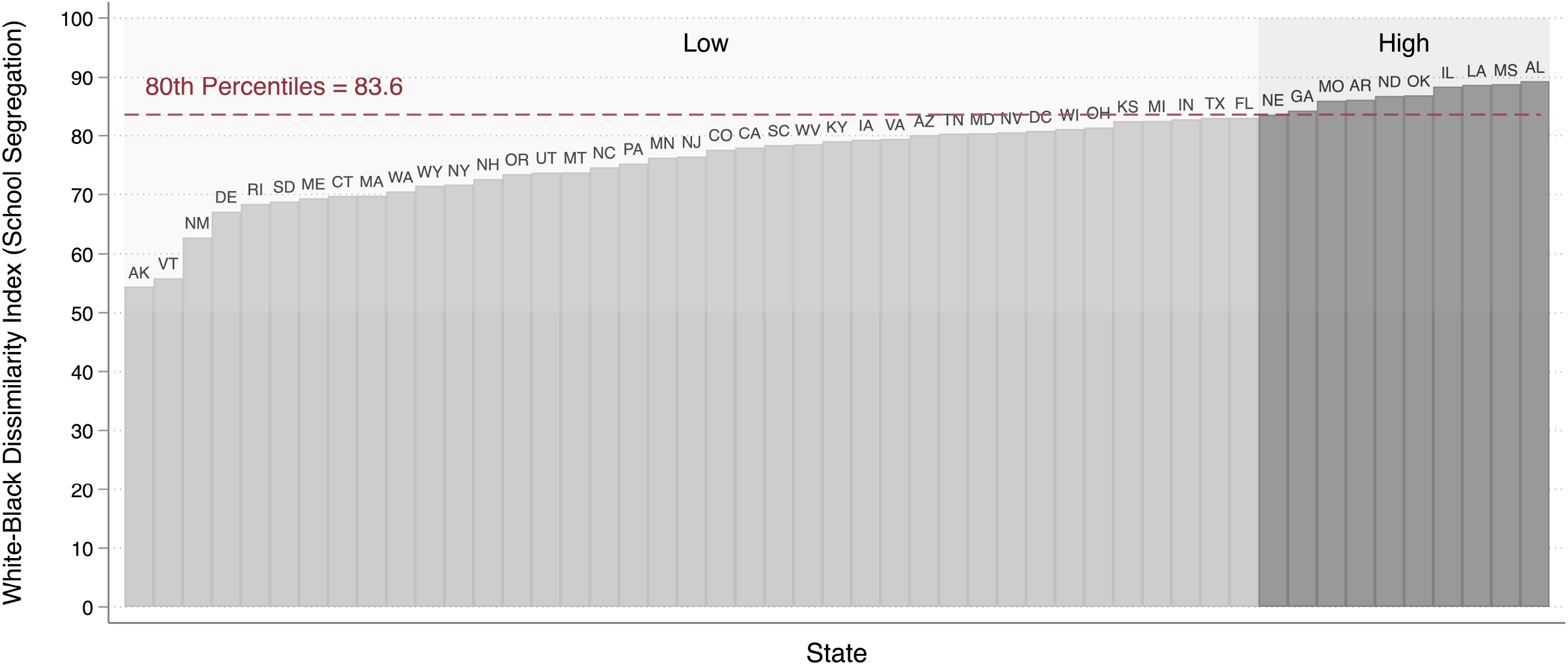
Distribution of school segregation at the state level *Notes*: The figure presents the levels of school segregation in late 1960s measured at the state level. The vertical bar denotes the White-Black dissimilarity index (representing the absolute levels of school segregation) for each state, and states are ordered by their index scores. Standard two-letter state abbreviations are displayed at the top of corresponding bars. States with dissimilarity index higher than 80^th^ percentiles (i.e., dissimilarity index ≥ 83.6) are categorized as “high segregation” (in dark gray color), and otherwise as “low segregation” (in light gray color). White refers to non-Hispanic White, and Black refers to non-Hispanic Black.

**Supplementary eFigure 2.**
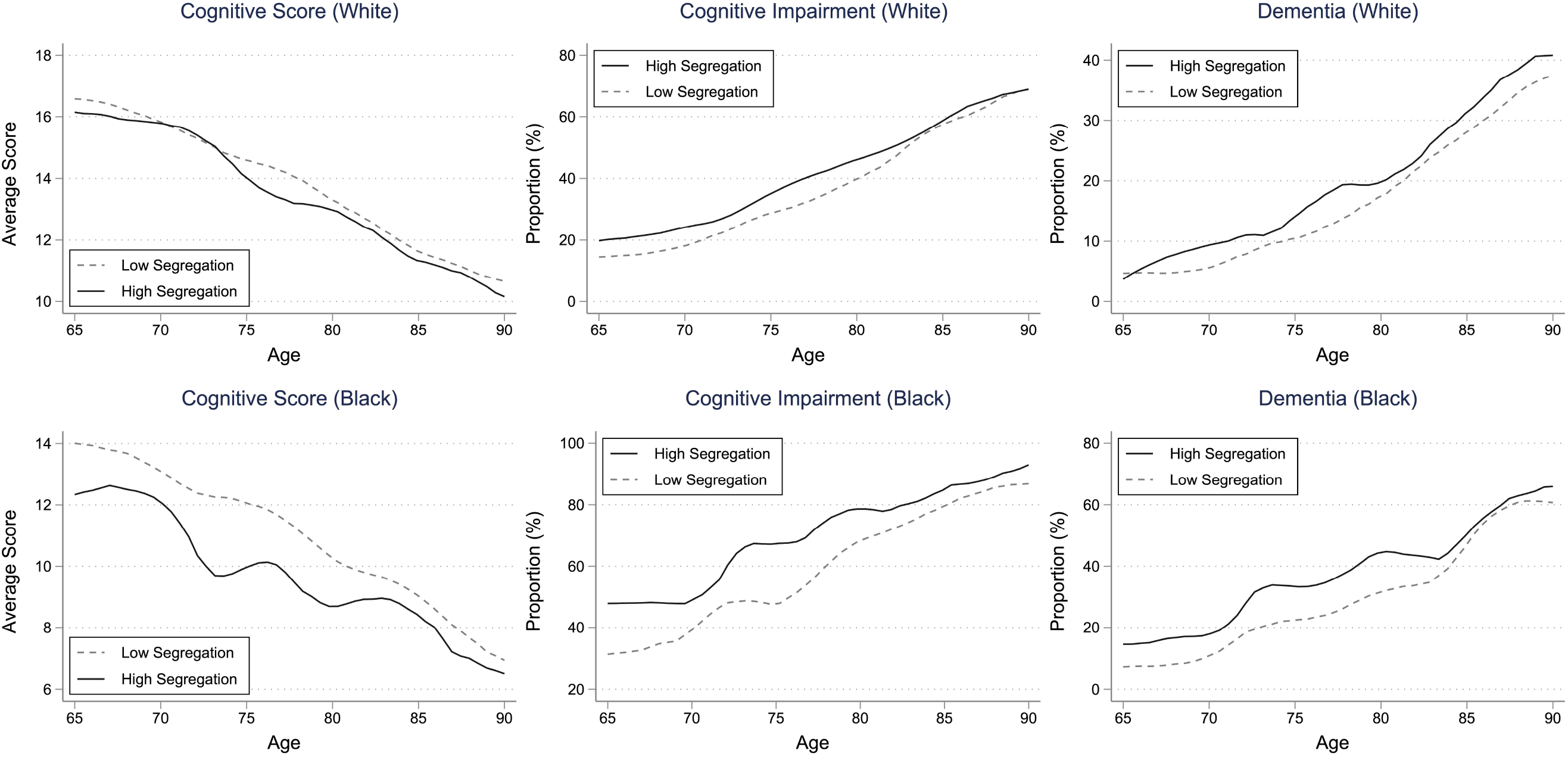
Trajectories of cognitive outcomes across ages for participants with high vs low levels of school segregation *Notes*: The figure presents the trajectories of cognitive score, cognitive impairment and dementia across age for White and Black participants. In each panel, sample were stratified into high vs. low levels of school segregation based on quintiles of White-Black dissimilarity index scores, and the trajectories for each group were fitted and plotted. The lines were fitted using local moving average. White refers to non-Hispanic White, and Black refers to non-Hispanic Black.

**Supplementary eTable 1.** Association between school segregation and cognitive outcomes for White and Black participants combined.

|  | (1) | (2) | (3) |
| --- | --- | --- | --- |
|  | All Participants |  |  |
| VARIABLES | Model A | Model B | Model C |
| **Panel A. Cognitive Score (0-27)** | *β* Coefficient (95% CI) |  |  |
| School Segregation | -0.71**<br>(-1.18, -0.24) | -0.32*<br>(-0.58, -0.06) | -0.29*<br>(-0.54, -0.04) |
| Observations | 16,420 | 16,420 | 16,420 |
| **Panel B. Cognitive Impairment (0/1)** | Adjusted Odds Ratio (95% CI) |  |  |
| School Segregation | 1.39**<br>(1.11, 1.74) | 1.24**<br>(1.06, 1.43) | 1.23**<br>(1.06, 1.42) |
| Observations | 19,960 | 19,960 | 19,960 |
| **Panel C. Dementia (0/1)** | Adjusted Odds Ratio (95% CI) |  |  |
| School Segregation | 1.35*<br>(1.04, 1.74) | 1.19*<br>(1.02, 1.38) | 1.17*<br>(1.02, 1.34) |
| Observations | 19,960 | 19,960 | 19,960 |
| Demographics | YES | YES | YES |
| SES | NO | YES | YES |
| Health | NO | NO | YES |

**Supplementary eTable 2.**
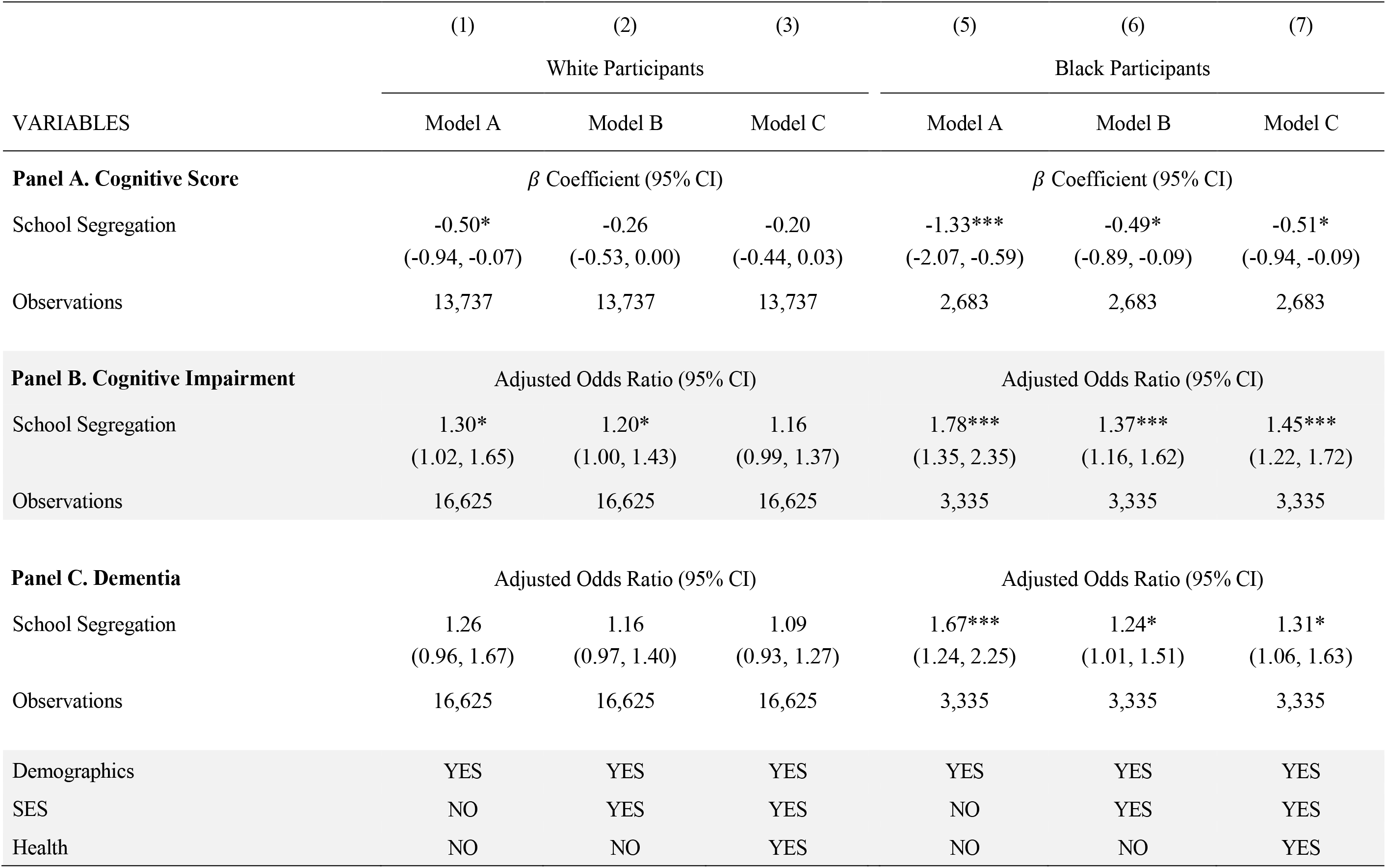
Association between school segregation and cognitive outcomes for White and Black participants, respectively.

|  | (1) | (2) | (3) | (5) | (6) | (7) |
| --- | --- | --- | --- | --- | --- | --- |
|  | White Participants |  |  | Black Participants |  |  |
| VARIABLES | Model A | Model B | Model C | Model A | Model B | Model C |
| **Panel A. Cognitive Score** | $\beta$ Coefficient (95% CI) | | | $\beta$ Coefficient (95% CI) | | |
| School Segregation | -0.50*<br>(-0.94, -0.07) | -0.26<br>(-0.53, 0.00) | -0.20<br>(-0.44, 0.03) | -1.33***<br>(-2.07, -0.59) | -0.49*<br>(-0.89, -0.09) | -0.51*<br>(-0.94, -0.09) |
| Observations | 13,737 | 13,737 | 13,737 | 2,683 | 2,683 | 2,683 |
| **Panel B. Cognitive Impairment** | Adjusted Odds Ratio (95% CI) |  |  | Adjusted Odds Ratio (95% CI) |  |  |
| School Segregation | 1.30*<br>(1.02, 1.65) | 1.20*<br>(1.00, 1.43) | 1.16<br>(0.99, 1.37) | 1.78***<br>(1.35, 2.35) | 1.37***<br>(1.16, 1.62) | 1.45***<br>(1.22, 1.72) |
| Observations | 16,625 | 16,625 | 16,625 | 3,335 | 3,335 | 3,335 |
| **Panel C. Dementia** | Adjusted Odds Ratio (95% CI) |  |  | Adjusted Odds Ratio (95% CI) |  |  |
| School Segregation | 1.26<br>(0.96, 1.67) | 1.16<br>(0.97, 1.40) | 1.09<br>(0.93, 1.27) | 1.67***<br>(1.24, 2.25) | 1.24*<br>(1.01, 1.51) | 1.31*<br>(1.06, 1.63) |
| Observations | 16,625 | 16,625 | 16,625 | 3,335 | 3,335 | 3,335 |
| Demographics | YES | YES | YES | YES | YES | YES |
| SES | NO | YES | YES | NO | YES | YES |
| Health | NO | NO | YES | NO | NO | YES |

